# Assessing psychosocial adversities at Cardiovascular-Kidney-Metabolic Syndrome stages in a large population survey

**DOI:** 10.64898/2026.09.22.26363738

**Authors:** Tanvir Ahmed, Darshan Chudasama, Biplab Datta, Marlo Vernon, Steven S. Coughlin, Jennifer C. Sullivan

## Abstract

**Background:** Cardiovascular-Kidney-Metabolic Syndrome (CKMS) is defined as a cluster of disease conditions that are highly prevalent in the U.S. population. This study aims to assess psychosocial adversities at different CKMS stages in a large survey of U.S. adults, measured by lack of emotional support, low life satisfaction, and loneliness.

**Methods:** This cross-sectional study used data on 159,511 adults from the 2023 Behavioral Risk Factor Surveillance System (BRFSS) survey. Proxy CKMS stages (0, 1, 2/3, and 4) were determined using self-reported health conditions. Separate multivariable logistic regression models were estimated to obtain the predictive margins of respective psychosocial attributes across proxy CKMS stages.

**Results:** Compared to adults at lower CKMS stages (Stages 0 and 1), adults at advanced CKMS stages (Stages 2/3 and 4) generally reported having higher psychosocial toll. At CKMS stage 4, the predicted probabilities of lack of emotional support, low life satisfaction, and loneliness were 6.96, 3.24, and 10.10 percentage points (pp) higher, respectively, than at stage 0. Compared to stage 1, the corresponding differences were 5.92, 2.89, and 9.65 pp. At CKMS stage 2/3, the corresponding differences were 4.36, 1.66, and 5.69 pp compared to stage 0 and 3.33, 1.32, and 5.25 pp compared to stage 1. The high burden at advanced CKMS stages was generally evident across sex, and different socioeconomic status conditions manifested by income and education.

**Conclusions:** Findings of this study suggest that adults with advanced CKMS stages were more likely to experience a psychosocial toll. Further research is warranted to facilitate strategies for improving psychosocial wellbeing of CKMS patients.

**What is Known:**

- Loneliness and social isolation have been associated with adverse cardiovascular outcomes.
- Prior research has examined loneliness and CKM stage together in relation to depression and anxiety, but evidence across other psychosocial outcomes remains limited.

**What the Study Adds:**

- Adults at advanced proxy CKMS stages had higher adjusted predicted probabilities of lacking emotional support, having low life satisfaction, and feeling lonely.
- These patterns were observed across sex, income, and education groups.

## Introduction

Cardiovascular-kidney-metabolic syndrome (CKMS) is a multi-system disorder characterized by cardiovascular disease, chronic kidney disease, and metabolic disorders such as obesity and diabetes, that are associated with increased morbidity and mortality.^1^ These interrelated conditions are among the leading causes of death in the U.S. and other countries. Recent studies conducted in the U.S. have found that almost 90% of adults meet criteria for CKMS stage 1 or higher, and that poor health due to CKMS is widespread in the population.^2–4^

Numerous previous studies have shown that social support is associated with improved health and lower risk of morbidity and mortality from cardiovascular diseases.^5,6^ Emotional support, which is an important part of social support, contributes to healthy adjustment to life.^5^ Loneliness, the discrepancy between a person’s desired and actual social relationships, is an emotional response to social isolation.^7^ Social isolation, on the other hand, is an objective measure of a lack of social corrections or interactions.^6,7^ Social isolation and loneliness are frequent sources of chronic stress in adults.^8^ Loneliness and social isolation have been associated with increased mortality.^8,9^ Social isolation also accelerates the onset of cardiovascular disease in persons with chronic kidney disease.^10^ A systematic review and meta-analysis of 16 prospective studies showed that loneliness and social isolation are associated with coronary heart disease and stroke.^11^ Few studies of loneliness in persons with CKMS have been reported. In a prospective study in the United Kingdom, Huang et al. found additive interactions between loneliness and cardiovascular-kidney-metabolic health on the risk of depression and anxiety. Participants who reported being lonely and in stage 4 CKMS had the greatest risk of depression and anxiety compared with those without loneliness and in stage 0.^12^

Life satisfaction, which is an important determinant of health, relates to the subjective wellbeing of how people evaluate the quality of their lives.^13,14^ In a national representative, cross-sectional study in China, Liu et al. found that life satisfaction is reduced in patients with cardiovascular disease who have an increased number of comorbidities or kidney disease.^14^ Studies of life satisfaction in patients with CKMS have not been previously reported.

In view of the importance of emotional support, loneliness, and life satisfaction in chronic illnesses such as cardiovascular disease and kidney disease, we intended to examine the prevalence of adverse psychosocial outcomes at different CKMS stages, in a large survey of adults in the U.S. Psychosocial toll was captured by three separate constructs – lack of emotional support, low life satisfaction, and loneliness. Note that in this cross-sectional analysis, we did not intend to examine whether CKMS conditions were the cause of any adverse psychosocial outcomes. Rather we intend to investigate whether the probability of experiencing adverse psychosocial outcomes varied at a given point of time across different CKMS stages. Since psychosocial wellbeing can play a critical role in health maintenance and disease progression, knowing the burden on psychosocial adversities across CKMS stages may help better CKMS management.

In the absence of clinical data to determine CKMS stages as per the American Heart Association guidelines, self-reported health conditions were systematically aligned with specific clinical conditions to determine proxy CKMS stages. This approach has been previously used to assess CKMS-related outcomes in population survey data.^15,16^ Utilizing this approach and taking advantage of available psychosocial wellbeing information in the population survey, we aimed to assess the likelihood of adverse psychosocial outcomes at different CKMS stages. Since psychosocial outcomes are closely related to socioeconomic status conditions,^17^ our secondary aim was to examine probable heterogeneities in the association between proxy CKMS stages and lack of emotional support, low life satisfaction, and loneliness across sex, income, and educational attainment.

## Methods

### Data

We used data from the 2023 Behavioral Risk Factor Surveillance System (BRFSS) survey. Our sample included 159,511 adults (aged 18+ years) residing in 40 states across the U.S. that administered the Social Determinants of Health and Health Equity module in the 2023 BRFSS. Observations for which health information to determine proxy CKMS stages, information about constructs of psychosocial wellbeing, and demographic and socioeconomic characteristics were missing, were excluded from analysis. A flow chart of the study sample is presented in Supplemental Figure S1. Note that we could not use 2024 BRFSS, the most recent survey year, since data on hypertension and high cholesterol were not available in the 2024 survey.

BRFSS data is publicly available and anonymized. The study was determined as “not human subjects research” by the Augusta University IRB.

### Measures

Proxy CKMS stages were determined by self-reported data on height and weight, self-reported diagnoses of diabetes, pre-diabetes, hypertension, high cholesterol, chronic kidney disease, and cardiovascular disease (myocardial infarction, coronary heart disease, and stroke). The algorithm of determining CKMS stages using self-reported health data is as previously reported by Coughlin et al. (2026). Due to data limitations, stages 2 and 3 were consolidated into a single stage. Our exposure variable thus consisted of 4 proxy CKMS categories – stage 0, stage 1, stage 2/3, and stage 4.

Lack of emotional support was defined as rarely, never, or sometimes getting needed social and emotional support as opposed to always or usually. Low life satisfaction was defined as dissatisfied or very dissatisfied with life as opposed to being very satisfied or satisfied. Loneliness was defined as always, usually, or sometimes feeling lonely as opposed to rarely or never.^18^ We thus have three separate binary outcome variables, denoting adverse psychosocial outcomes.

### Statistical analysis

For each psychosocial construct, separate multivariable logistic regression was estimated to calculate predictive margins at proxy CKMS stages. Adjustments were made for age, age squared (to account for non-linear influence of age on both exposure and outcome), sex, race and ethnicity, marital status, income (as ratio of Federal Poverty Level [FPL]), educational attainment, employment status, urban/rural residence, number of household members (family size), health insurance, interview month fixed effect, and state of resident fixed effect. Adjusted odds ratios from the three multivariable logistic regression models are presented in Supplemental Table S4.

Categories of proxy CKMS stages were separately interacted with sex (male and female), income (low, middle, and high), and education (≤ high school, some college, and college graduate) to assess potential heterogeneities. Low, middle, and high income are defined as income less than 200% of the FPL, 200% to less than 400% of the FPL, and greater or equals 400% of the FPL, respectively.

Analyses were performed in Stata 18.0 software. The complex survey weights entailing the sampling frame of the BRFSS were used while conducting analyses.

## Results

Weighted prevalence of three psychosocial outcomes and the weighted distribution of participant’s sociodemographic and socioeconomic contributions across the proxy CKMS stages are presented in Table 1. Among 159,511 participants, 15.15% were determined to be at CKMS stage 0, and 26.67%, 50.1%, and 8.07% were determined to be at CKMS stages 1, 2/3, and 4, respectively.

**Table 1.**
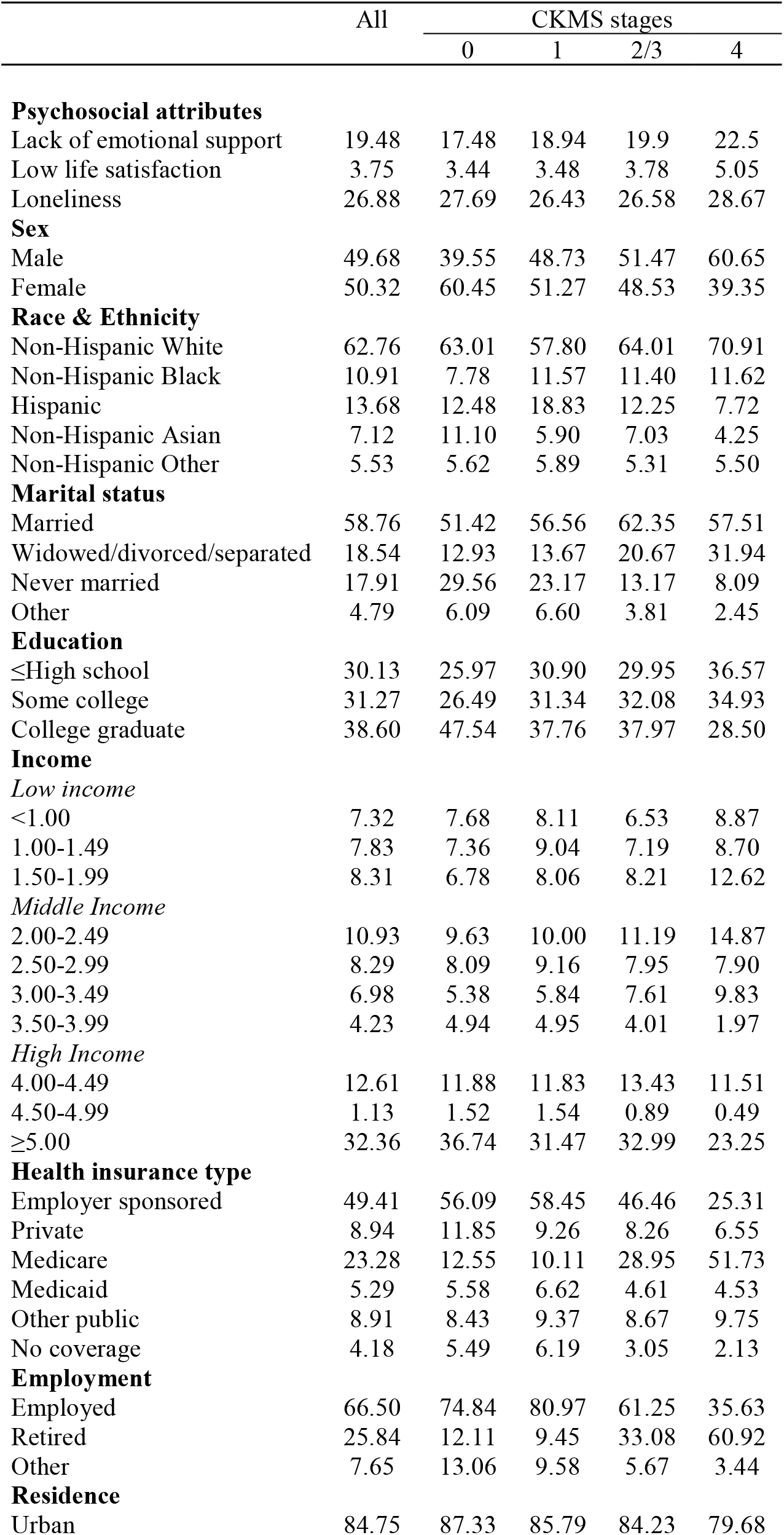
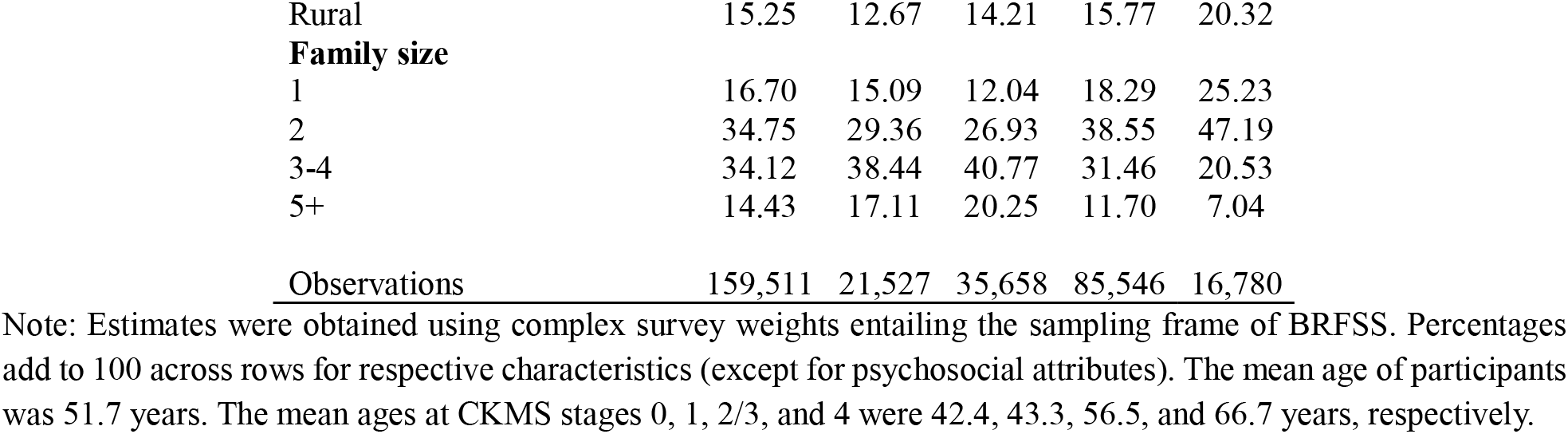
Prevalence (%) of psychosocial attributes and distribution (%) of study sample by proxy CKMS stages.

|  | All | CKMS stages |  |  |  |
| --- | --- | --- | --- | --- | --- |
|  |  | 0 | 1 | 2/3 | 4 |
| **Psychosocial attributes** |  |  |  |  |  |
| Lack of emotional support | 19.48 | 17.48 | 18.94 | 19.9 | 22.5 |
| Low life satisfaction | 3.75 | 3.44 | 3.48 | 3.78 | 5.05 |
| Loneliness | 26.88 | 27.69 | 26.43 | 26.58 | 28.67 |
| **Sex** |  |  |  |  |  |
| Male | 49.68 | 39.55 | 48.73 | 51.47 | 60.65 |
| Female | 50.32 | 60.45 | 51.27 | 48.53 | 39.35 |
| **Race & Ethnicity** |  |  |  |  |  |
| Non-Hispanic White | 62.76 | 63.01 | 57.80 | 64.01 | 70.91 |
| Non-Hispanic Black | 10.91 | 7.78 | 11.57 | 11.40 | 11.62 |
| Hispanic | 13.68 | 12.48 | 18.83 | 12.25 | 7.72 |
| Non-Hispanic Asian | 7.12 | 11.10 | 5.90 | 7.03 | 4.25 |
| Non-Hispanic Other | 5.53 | 5.62 | 5.89 | 5.31 | 5.50 |
| **Marital status** |  |  |  |  |  |
| Married | 58.76 | 51.42 | 56.56 | 62.35 | 57.51 |
| Widowed/divorced/separated | 18.54 | 12.93 | 13.67 | 20.67 | 31.94 |
| Never married | 17.91 | 29.56 | 23.17 | 13.17 | 8.09 |
| Other | 4.79 | 6.09 | 6.60 | 3.81 | 2.45 |
| **Education** |  |  |  |  |  |
| ≤High school | 30.13 | 25.97 | 30.90 | 29.95 | 36.57 |
| Some college | 31.27 | 26.49 | 31.34 | 32.08 | 34.93 |
| College graduate | 38.60 | 47.54 | 37.76 | 37.97 | 28.50 |
| **Income** |  |  |  |  |  |
| *Low income* |  |  |  |  |  |
| <1.00 | 7.32 | 7.68 | 8.11 | 6.53 | 8.87 |
| 1.00-1.49 | 7.83 | 7.36 | 9.04 | 7.19 | 8.70 |
| 1.50-1.99 | 8.31 | 6.78 | 8.06 | 8.21 | 12.62 |
| *Middle Income* |  |  |  |  |  |
| 2.00-2.49 | 10.93 | 9.63 | 10.00 | 11.19 | 14.87 |
| 2.50-2.99 | 8.29 | 8.09 | 9.16 | 7.95 | 7.90 |
| 3.00-3.49 | 6.98 | 5.38 | 5.84 | 7.61 | 9.83 |
| 3.50-3.99 | 4.23 | 4.94 | 4.95 | 4.01 | 1.97 |
| *High Income* |  |  |  |  |  |
| 4.00-4.49 | 12.61 | 11.88 | 11.83 | 13.43 | 11.51 |
| 4.50-4.99 | 1.13 | 1.52 | 1.54 | 0.89 | 0.49 |
| ≥5.00 | 32.36 | 36.74 | 31.47 | 32.99 | 23.25 |
| **Health insurance type** |  |  |  |  |  |
| Employer sponsored | 49.41 | 56.09 | 58.45 | 46.46 | 25.31 |
| Private | 8.94 | 11.85 | 9.26 | 8.26 | 6.55 |
| Medicare | 23.28 | 12.55 | 10.11 | 28.95 | 51.73 |
| Medicaid | 5.29 | 5.58 | 6.62 | 4.61 | 4.53 |
| Other public | 8.91 | 8.43 | 9.37 | 8.67 | 9.75 |
| No coverage | 4.18 | 5.49 | 6.19 | 3.05 | 2.13 |
| **Employment** |  |  |  |  |  |
| Employed | 66.50 | 74.84 | 80.97 | 61.25 | 35.63 |
| Retired | 25.84 | 12.11 | 9.45 | 33.08 | 60.92 |
| Other | 7.65 | 13.06 | 9.58 | 5.67 | 3.44 |
| **Residence** |  |  |  |  |  |
| Urban | 84.75 | 87.33 | 85.79 | 84.23 | 79.68 |
| Rural | 15.25 | 12.67 | 14.21 | 15.77 | 20.32 |
| **Family size** |  |  |  |  |  |
| 1 | 16.70 | 15.09 | 12.04 | 18.29 | 25.23 |
| 2 | 34.75 | 29.36 | 26.93 | 38.55 | 47.19 |
| 3-4 | 34.12 | 38.44 | 40.77 | 31.46 | 20.53 |
| 5+ | 14.43 | 17.11 | 20.25 | 11.70 | 7.04 |
| <u>Observations</u> | <u>159,511</u> | <u>21,527</u> | <u>35,658</u> | <u>85,546</u> | <u>16,780</u> |

Note: Estimates were obtained using complex survey weights entailing the sampling frame of BRFSS. Percentages add to 100 across rows for respective characteristics (except for psychosocial attributes). The mean age of participants was 51.7 years. The mean ages at CKMS stages 0, 1, 2/3, and 4 were 42.4, 43.3, 56.5, and 66.7 years, respectively.

The mean age of participants was 51.7 years. The mean age was lower at CKMS stages 0 and 1 (42.4 and 43.3 years, respectively), and higher at CKMS stages 2/3 and 4 (56.5 and 66.7 years, respectively). While sex composition was near-even at stages 1 and 2/3, females were predominant at stage 0 (60.45%), and males were predominant at stage 4 (60.65%). A higher share of non-Hispanic Whites, and lower shares of Hispanic and non-Hispanic Asian were observed at stage 4. Share of college graduates was relatively higher at CKMS stage 0, and lower at CKMS stage 4. Conversely, share of adults with educational attainment of high school or less was lower at CKMS stage 0 and higher at stage 4. Share of the highest income band (i.e., ≥ 500% of FPL) was lower at CKMS stage 4, whereas the distribution of adults at the lowest income band (i.e., < 100% of FPL) was similar across CKMS stages.

### Lack of emotional support

Prevalence of lack of emotional support was 19.48% in the study sample. While it was 17.48% among adults at CKMS stage 0, the prevalence was 5.02 percentage points (95% CI: 3.23 – 6.80) higher among those at CKMS stage 4. Adjusted predicted probability of lack of emotional support at different CKMS stages are presented in Figure 1. Compared to predicted probabilities (multiplied by 100 and expressed as %) of lack of emotional support at CKMS stages 0 and 1 (16.53% and 17.57%, respectively), probabilities at stages 2/3 and 4 were significantly higher (20.89% and 23.59%, respectively [p<0.001]). The higher predicted probability of lack of emotional support at advanced CKMS stages was generally evident across sex, educational attainment, and income groups. Crude prevalence of lack of emotional support was higher among males, those with low family income, and those with a high school education or less (Supplemental Table S1). Nevertheless, within each sex, income, and education subgroup, the adjusted predicted probability of lacking emotional support was higher at advanced CKMS stages (Figure 1).

**Figure 1.**
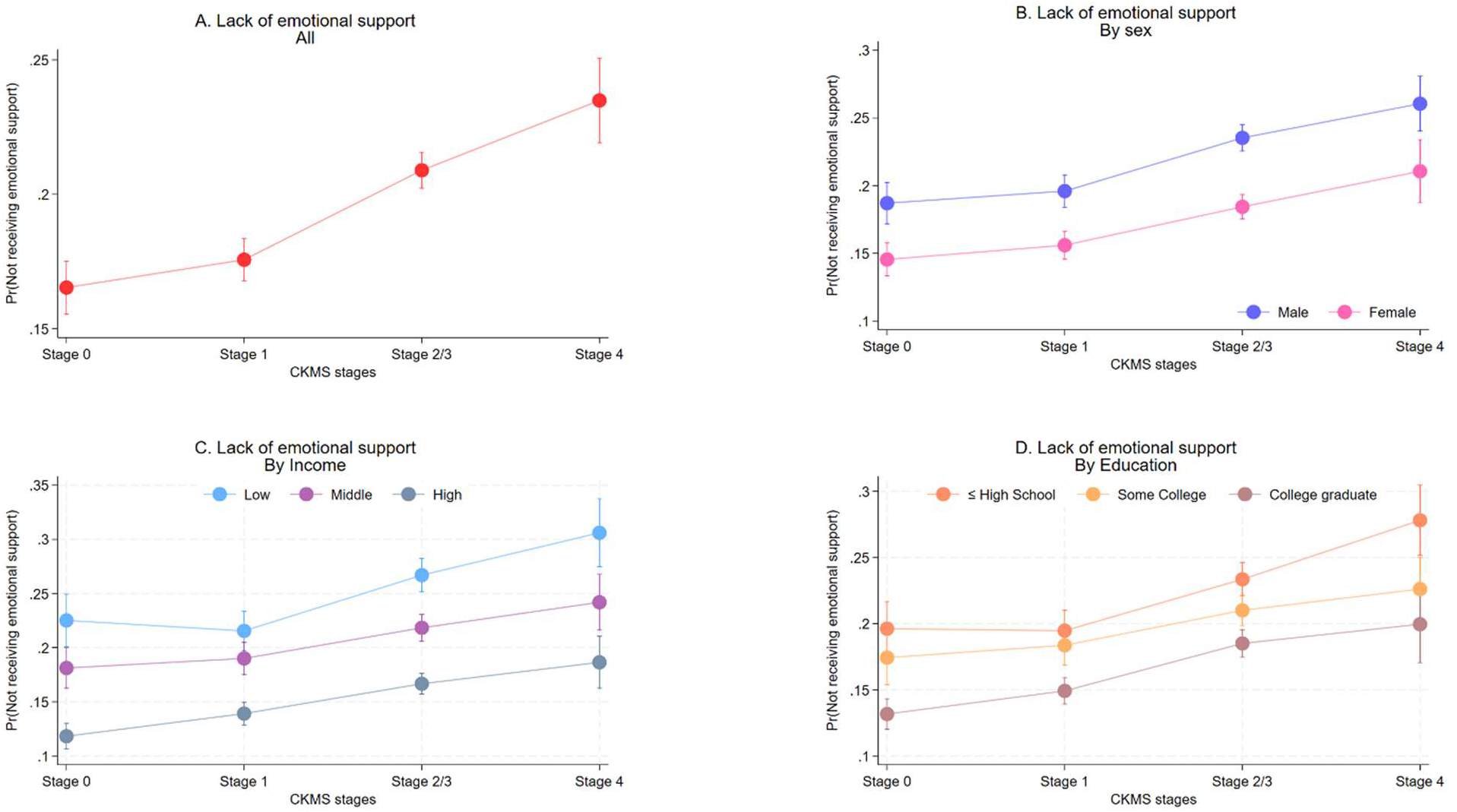
Predictive probabilities of lack of emotional support at proxy CKMS stages. Note: Models were estimated using complex survey weights entailing the sampling frame of the BRFSS. Models were adjusted for age, age squared, sex, race and ethnicity, marital status, income, educational attainment, health insurance coverage type, employment status, urban/rural residence, number of household members, interview month fixed effect, and state of resident fixed effect. Panels A–D show results without interaction, and interaction of emotional support indicator with sex, income group, and education group, respectively. Abbreviations: BRFSS, Behavioral Risk Factor Surveillance System; CKMS, cardiovascular-kidney-metabolic syndrome; Pr, probability.

### Low life satisfaction

3.75% of adults in the sample reported low life satisfaction. The prevalence was similar (ranging from 3.44% and 3.78%) across CKMS stages 0, 1, and 2/3, but significantly higher at stage 4 (5.05%). Adjusted predicted probability of low life satisfaction was 2.69% and 3.03% for adults at CKMS stages 0 and 1, respectively. For adults at CKMS stage 2/3 the probability increased to 4.36% and further increased to 5.93% for adults at stage 4 (Figure 2). Crude prevalence of low life satisfaction was higher among adults with low income and lower educational attainment, though no significant differences by sex were observed (Supplemental Table S2). Despite these general differences, adjusted likelihood of low life satisfaction was found higher at advanced CKMS stage across sex, income, and education (Figure 2).

**Figure 2.**
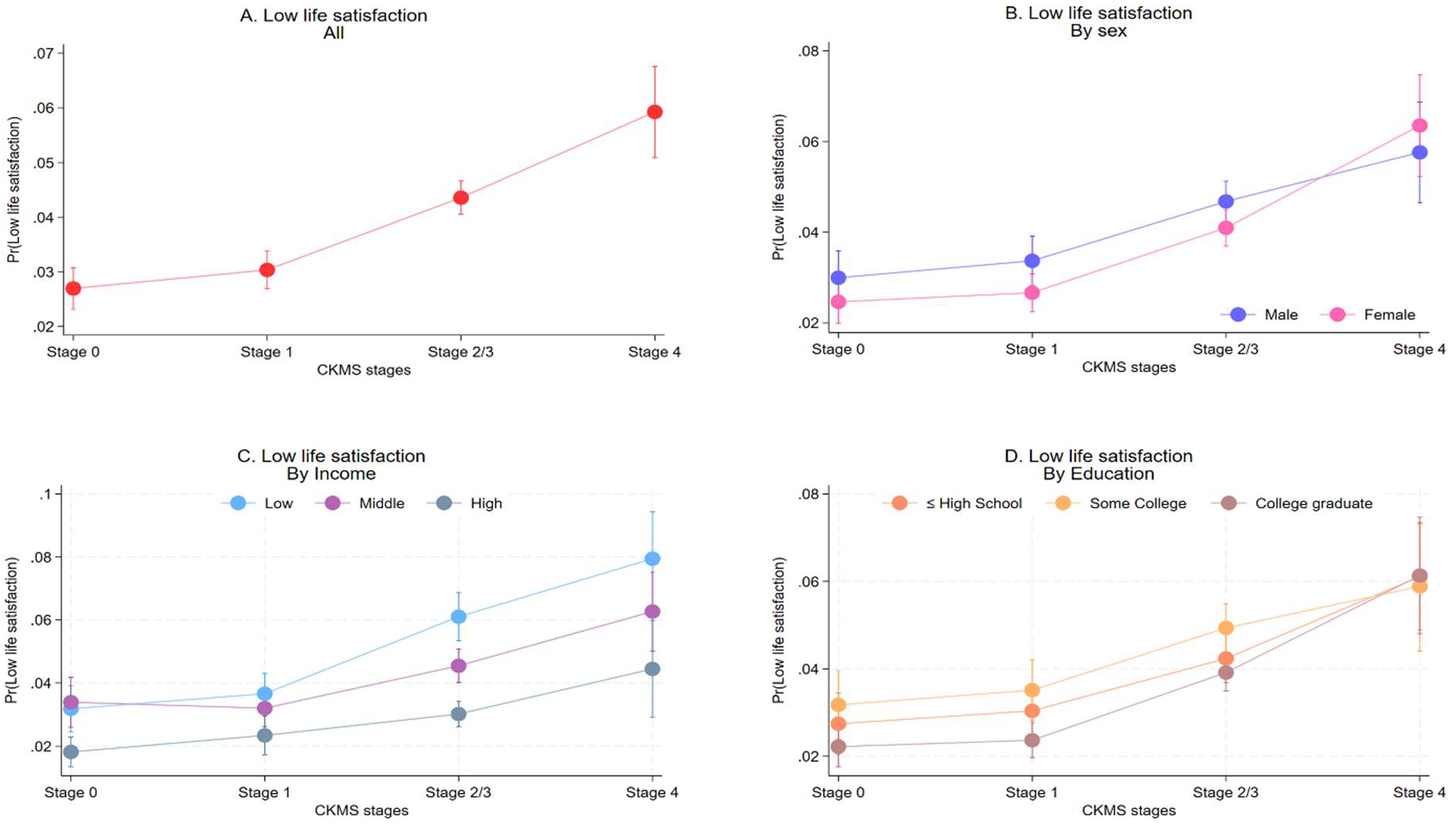
Predictive probabilities of low life satisfaction at proxy CKMS stages. Note: Models were estimated using complex survey weights entailing the sampling frame of the BRFSS. Models were adjusted for age, age squared, sex, race and ethnicity, marital status, income, educational attainment, health insurance coverage type, employment status, urban/rural residence, number of household members, interview month fixed effect, and state of resident fixed effect. Panels A–D show results without interaction, and interaction of life satisfaction indicator with sex, income group, and education group, respectively. Abbreviations: BRFSS, Behavioral Risk Factor Surveillance System; CKMS, cardiovascular-kidney-metabolic syndrome; Pr, probability.

### Loneliness

Prevalence of loneliness in the study sample was 26.88%. No significant differences in crude prevalence of loneliness were observed at different CKMS stages. However, after adjusting for demographic and socioeconomic covariates, significant higher likelihood of loneliness was observed at advanced CKMS stages, compared that at stages 0 and 1 (Figure 3). While crude prevalence of loneliness was lower among males and adults with higher income and educational attainment (Supplemental Table S3), the higher adjusted probability of loneliness at CKMS stages 2/3 and 4 was evident in both males and females and in all income and educational levels (Figure 3).

**Figure 3.**
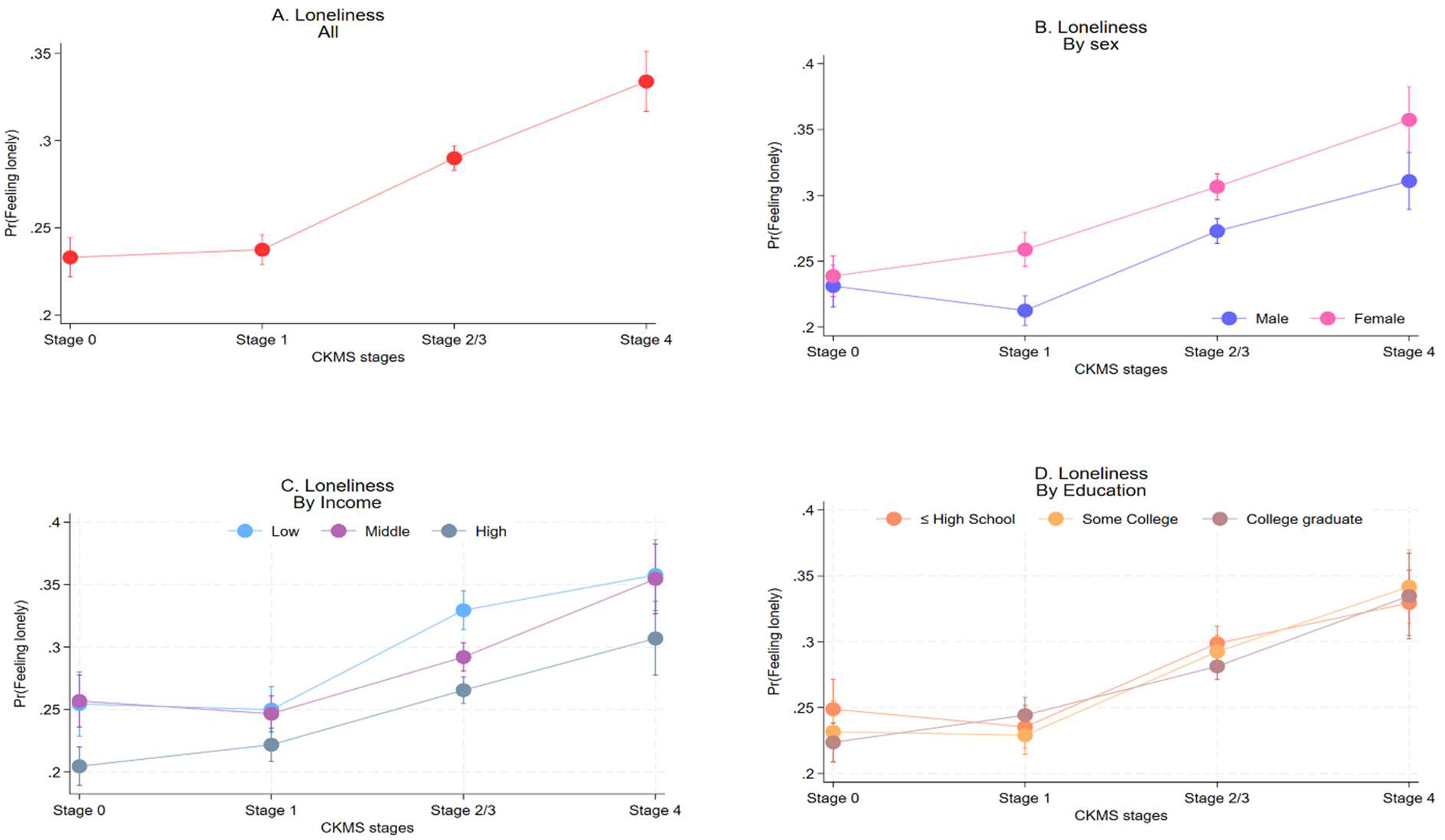
Predictive probabilities of loneliness at proxy CKMS stages. Note: Models were estimated using complex survey weights entailing the sampling frame of the BRFSS. Models were adjusted for age, age squared, sex, race and ethnicity, marital status, income, educational attainment, employment status, health insurance coverage type, urban/rural residence, number of household members, interview month fixed effect, and state of resident fixed effect. Panels A–D show results f without interaction, and interaction of loneliness indicator with sex, income group, and education group, respectively. Abbreviations: BRFSS, Behavioral Risk Factor Surveillance System; CKMS, cardiovascular-kidney-metabolic syndrome; Pr, probability.

## Discussion

In this large sample of 159,511 U.S. adults, living in 40 U.S. states, we found a consistent association between proxy CKMS stage and psychosocial adversity: adults at advanced CKMS stages (2/3 and 4) had a higher predicted likelihood of lacking emotional support, reporting low life satisfaction, and feeling lonely than their counterparts at CKMS stage 0 or stage 1. The pattern was broadly consistent across sex, income, and education subgroups, suggesting that the burden of psychosocial adversity at advanced CKMS stages is not confined to any single demographic or socioeconomic group, though the magnitude may differ across groups.

These findings extend a growing literature linking individual components of CKMS and psychosocial harm. Prior work has shown that social isolation accelerates the onset of cardiovascular disease among adults with chronic kidney disease.^10^ Loneliness and social isolation are also found to be associated with coronary heart disease and stroke.^11^ Evidence of reduced life satisfaction has been observed among adults with cardiovascular disease who carry additional comorbidities or kidney disease.^14^ Most directly relevant, Huang et al. reported that loneliness and CKM health interact additively to raise the risk of depression and anxiety, with the highest risk observed among adults who were both lonely and at CKMS stage 4.^12^

Our results are consistent with these findings and advances the literature by showing that CKMS stages are independently associated with a broader set of psychosocial constructs including emotional support, life satisfaction and loneliness. Another contribution of our analysis is that the findings are more generalizable as we used a larger US representative survey weighted study sample across sex, income, and education groups compared to prior CKM-focused studies on psychosocial wellbeing. Because CKMS stage is defined by the cluster of cardiovascular, kidney, and metabolic conditions rather than a single disease condition, our findings suggest that psychosocial burden increases with the cumulative physiological burden of CKM conditions, echoing the broader multimorbidity literature in which psychological distress increases as chronic conditions accumulate.^19,20^

While we were unable to evaluate any inter-temporal association, we offer some probable explanations for observing the higher likelihood of psychosocial adversities at higher CKMS stages, which may be empirically examined in future studies utilizing appropriate data. First, chronic stress that arises from multiple concurrent conditions, more frequent healthcare visits, more complex medication regimens, and greater functional limitation, may itself reduce social participation and subjective wellbeing.^21,22^ Second, physical symptoms and disability associated with late-stage cardiovascular, kidney, and metabolic disease may directly limit the ability to maintain social relationships or engage in activities that support life satisfaction.^14,22^ Third, the association could be bidirectional, that is psychosocial factors may not only result from CKMS conditions but may also influence it.^11,14^

### Strengths and Limitations

This study has several strengths. It draws on a large sample with complex survey weighting, uses a previously described algorithm for staging proxy CKMS from self-reported health data (Coughlin et al.), examines three distinct and clinically meaningful psychosocial constructs, and includes stratified analyses that showed the consistency of the CKMS-psychosocial gradient across sex, income, and education.^4^

Several limitations should also be noted. First, the cross-sectional design precludes causal inference and cannot establish any temporal or bidirectional relationship between CKMS progression and burden of psychosocial adversities. Second, both CKMS stage and the psychosocial outcomes were based on self-report, which may introduce misclassification. In particular, the proxy algorithm collapses CKMS stages 2 and 3 because BRFSS lacks measures of kidney damage and subclinical cardiovascular disease. Third, emotional support, life satisfaction, and loneliness were each assessed with single-item measures rather than longer validated instruments (e.g., the UCLA Loneliness Scale), and the response thresholds differed across outcomes, which may limit measurement precision and comparability across the three constructs.^23^ Fourth, although our models adjusted for a broad set of demographic and socioeconomic covariates, residual confounding from unmeasured factors cannot be entirely ruled out.

## Conclusions

Notwithstanding the limitations, our findings demonstrate higher adjusted predicted probabilities of psychosocial adversities at advanced CKMS stages. These findings provide new avenues for research to facilitate better management of CKMS conditions. For example, current CKMS staging and management frameworks primarily focus on cardiovascular, renal, and metabolic risk reduction. Building on our findings, future research may explore opportunities for psychosocial screening to be incorporated into care for adults across CKMS stages, and potential of community-based social support resources for efficient CKMS management.

## Data Availability

Data used in this study is publicly available from the Centers for Disease Control and Prevention (CDC) website.

https://www.cdc.gov/brfss/annual_data/annual_2024.html

## Acknowledgements

N/A.

## Source of Funding

Funding for this research was provided by the American Heart Association (grants nos. 25SFRNCCKMS1442888 and 25SFRNPCKMS1467445).

## Disclosures

None.

## Notes

### Competing Interest Statement

The authors have declared no competing interest.

### Clinical Trial

This study is not a clinical trial.

### Author Declarations

This study was determined "Not Human Subjects Research" by the Augusta University Institutional Review Board.

## References

1. Ndumele CE, Rangaswami J, Chow SL, Neeland IJ, Tuttle KR, Khan SS, Coresh J, Mathew RO, Baker-Smith CM, Carnethon MR, et al. Cardiovascular-Kidney-Metabolic Health: A Presidential Advisory From the American Heart Association. Circulation. 2023;148:1606– 1635.

2. Minhas AMK, Mathew RO, Sperling LS, Nambi V, Virani SS, Navaneethan SD, Shapiro MD, Abramov D. Prevalence of the Cardiovascular-Kidney-Metabolic Syndrome in the United States. J. Am. Coll. Cardiol. 2024;83:1824–1826.

3. Aggarwal R, Ostrominski JW, Vaduganathan M. Prevalence of Cardiovascular-Kidney-Metabolic Syndrome Stages in US Adults, 2011-2020. JAMA. 2024;331:1858–1860.

4. Coughlin SS, Parikh N, Oh A, Datta B, Vernon M, Sullivan J. The Prevalence of Cardiovascular–Kidney–Metabolic Syndrome: A Review of Published Estimates and New Findings from BRFSS Surveys. Cardiovasc. Med. 2026;29:5.

5. Zhu Y, Que D, Jin Z, Zhang X, Song X, Chen K, Yang P. Association of different emotional support status with cardio-cerebrovascular diseases. J. Affect. Disord. 2025;374:303–311.

6. Cené CW, Beckie TM, Sims M, Suglia SF, Aggarwal B, Moise N, Jiménez MC, Gaye B, McCullough LD, the American Heart Association Social Determinants of Health Committee of the Council on Epidemiology and Prevention and Council on Quality of Care and Outcomes Research; Prevention Science Committee of the Council on Epidemiology and Prevention and Council on Cardiovascular and Stroke Nursing; Council on Arteriosclerosis, Thrombosis and Vascular Biology; and Stroke Council. Effects of Objective and Perceived Social Isolation on Cardiovascular and Brain Health: A Scientific Statement From the American Heart Association. J. Am. Heart Assoc. 2022;11:e026493.

7. Xia N, Li H. Loneliness, Social Isolation, and Cardiovascular Health. Antioxid. Redox Signal. 2018;28:837–851.

8. Steptoe A, Shankar A, Demakakos P, Wardle J. Social isolation, loneliness, and all-cause mortality in older men and women. Proc. Natl. Acad. Sci. 2013;110:5797–5801.

9. Holt-Lunstad J, Smith TB, Baker M, Harris T, Stephenson D. Loneliness and Social Isolation as Risk Factors for Mortality: A Meta-Analytic Review. Perspect. Psychol. Sci. 2015;10:227–237.

10. Zeng X, Jiang Y, Liu Z, Yang H, Song H, Li C, Fu P. Social Isolation Is Associated With the Acceleration of Death and Incident Cardiovascular Disease in Adults With Chronic Kidney Disease. J. Am. Heart Assoc. 2025;14:e038951.

11. Valtorta NK, Kanaan M, Gilbody S, Ronzi S, Hanratty B. Loneliness and social isolation as risk factors for coronary heart disease and stroke: systematic review and meta-analysis of longitudinal observational studies. Heart. 2016;102:1009–1016.

12. Huang X, Liang J, Zhang J, Fu J, Xie W, Zheng F. Association of cardiovascular-kidney-metabolic health and social connection with the risk of depression and anxiety. Psychol. Med. 2024;54:4203–4211.

13. Shad B, Ashouri A, Hasandokht T, Rajati F, Salari A, Naghshbandi M, Mirbolouk F. Effect of multimorbidity on quality of life in adult with cardiovascular disease: a cross-sectional study. Health Qual. Life Outcomes. 2017;15:240.

14. Liu G, Xue Y, Liu Y, Wang S, Geng Q. Multimorbidity in cardiovascular disease and association with life satisfaction: a Chinese national cross-sectional study. BMJ Open. 2020;10:e042950.

15. Sharma M, Chudasama D, Datta BK, Coughlin SS, Vernon MM, Sullivan JC. Cardiovascular-Kidney-Metabolic Syndrome and Health Risk Behaviors in U.S. Adults. AJPM Focus. 2026;100529.

16. Chang R, Parekh T, Hagan KK, Javed Z, Ostrominski JW. State-Level Prevalence of Cardiovascular-Kidney-Metabolic Syndrome Stages in the United States, 2011 to 2023. JACC Adv. 2025;4:101754.

17. Navarro-Carrillo G, Alonso-Ferres M, Moya M, Valor-Segura I. Socioeconomic Status and Psychological Well-Being: Revisiting the Role of Subjective Socioeconomic Status. Front. Psychol. [Internet]. 2020 [cited 2026 Sept 18];11. Available from: https://www.frontiersin.org/journals/psychology/articles/10.3389/fpsyg.2020.01303/full

18. Town M. Racial and Ethnic Differences in Social Determinants of Health and Health-Related Social Needs Among Adults — Behavioral Risk Factor Surveillance System, United States, 2022. MMWR Morb. Mortal. Wkly. Rep. [Internet]. 2024 [cited 2026 Sept 18];73. Available from: https://www.cdc.gov/mmwr/volumes/73/wr/mm7309a3.htm

19. Read JR, Sharpe L, Modini M, Dear BF. Multimorbidity and depression: A systematic review and meta-analysis. J. Affect. Disord. 2017;221:36–46.

20. Adzrago D, Williams DR, Williams F. Multiple chronic diseases and psychological distress among adults in the United States: the intersectionality of chronic diseases, race/ethnicity, immigration, sex, and insurance coverage. Soc. Psychiatry Psychiatr. Epidemiol. 2025;60:181–199.

21. Steptoe A, Kivimäki M. Stress and cardiovascular disease: an update on current knowledge. Annu. Rev. Public Health. 2013;34:337–354.

22. Fingerman KL, Ng YT, Huo M, Birditt KS, Charles ST, Zarit S. Functional Limitations, Social Integration, and Daily Activities in Late Life. J. Gerontol. Ser. B. 2021;76:1937–1947.

23. Russell DW. UCLA Loneliness Scale (Version 3): Reliability, Validity, and Factor Structure. J. Pers. Assess. 1996;66:20–40.

